# Plasma glial fibrillary acidic protein and neurofilament light chain in behavioural variant frontotemporal dementia and primary psychiatric disorders

**DOI:** 10.1101/2024.08.08.24311409

**Authors:** Dhamidhu Eratne, Matthew JY Kang, Courtney Lewis, Christa Dang, Charles Malpas, Suyi Ooi, Amy Brodtmann, David Darby, Henrik Zetterberg, Kaj Blennow, Michael Berk, Olivia Dean, Chad Bousman, Naveen Thomas, Ian Everall, Chris Pantelis, Cassandra Wannan, Claudia Cicognola, Oskar Hansson, Shorena Janelidze, Alexander F Santillo, Dennis Velakoulis, The MiND Study Group

**Affiliations:** Neuropsychiatry, Royal Melbourne Hospital, 300 Grattan St Parkville VIC 3052 Australia; Department of Psychiatry, University of Melbourne, Grattan St Parkville VIC 3052, Melbourne, Australia; The Florey Institute, 30 Royal Parade, Parkville VIC 3052, Australia; National Ageing Research Institute, 34-54 Poplar Rd, Parkville VIC 3052 Australia; Department of General Practice, University of Melbourne, Grattan St Parkville VIC 3052, Melbourne, Australia; Department of Medicine, Royal Melbourne Hospital, Grattan St Parkville VIC 3052, Melbourne, Australia; University of Melbourne, Grattan St Parkville VIC 3052, Melbourne, Australia; School of Translational Medicine, Monash University, Melbourne, Australia; Department of Neurology, RMH, 3050; Eastern Cognitive Disorders Clinic, Box Hill Hospital, Melbourne, Australia; Department of Psychiatry and Neurochemistry, Institute of Neuroscience and Physiology, the Sahlgrenska Academy at the University of Gothenburg, Universitetsplatsen 1, 405 30, Sweden; Clinical Neurochemistry Laboratory, Sahlgrenska University Hospital, Mölndal, SE-43180, Sweden; Department of Neurodegenerative Disease, UCL Institute of Neurology, Queen Square, London WC1N 3BG, UK; UK Dementia Research Institute at UCL, London WC1N 3BG, UK; Institute for Mental and Physical Health and Clinical Translation (IMPACT), Deakin University, Geelong, Australia; Department of Medical Genetics, University of Calgary, Calgary, AB, Canada; Sunshine Hospital, Melbourne, Australia; King’s College London, London, UK; Western Centre for Health Research & Education, University of Melbourne & Western Health, Sunshine Hospital, St Albans, Victoria, Australia; Monash Institute of Pharmaceutical Sciences (MIPS), Monash University, Parkville, Melbourne, Victoria, Australia; Centre for Youth Mental Health, The University of Melbourne, Parkville, VIC, Australia; Orygen, Parkville, VIC, Australia; Clinical Memory Research Unit, Department of Clinical Sciences Malmö, Faculty of Medicine, Lund University, Lund, Sweden

**Author notes:** Corresponding author: Dr Dhamidhu Eratne BHB MBChB FRANZCP, Neuropsychiatry, Royal Melbourne Hospital, 300 Grattan St, Parkville VIC 3050, Australia.

## Abstract

**Objective:** Timely, accurate distinction between behavioural variant frontotemporal dementia (bvFTD) and primary psychiatric disorders (PPD) is a clinical challenge. Blood biomarkers such as neurofilament light chain (NfL) and glial fibrillary acidic protein (GFAP) have shown promise. Prior work has shown NfL helps distinguish FTD from PPD. Few studies have assessed NfL together with GFAP.

**Methods:** We investigated plasma GFAP and NfL levels in participants with bvFTD, bipolar affective disorder (BPAD), major depressive disorder (MDD), treatment-resistant schizophrenia (TRS), healthy controls (HC), adjusting for age and sex. We compared ability of GFAP and NfL to distinguish bvFTD from PPD.

**Results:** Plasma GFAP levels were significantly (all p<0.001) elevated in bvFTD (n=22, mean (M)=273pg/mL) compared to BPAD (n=121, M=96pg/mL), MDD (n=42, M=105pg/mL), TRS (n=82, M=67.9pg/mL), and HC (n=120, M=76.8pg/mL). GFAP distinguished bvFTD from all PPD with an area under the curve (AUC) of 0.85, 95% confidence interval [0.76, 0.95]. The optimal cut-off of 105pg/mL was associated with 73% specificity and 86% sensitivity. NfL had AUC 0.95 [0.91, 0.99], 13.3pg/mL cut-off, 88% specificity, 86% sensitivity, and was superior to GFAP (p=0.02863) and combination of GFAP and NfL (p=0.04726).

**Conclusions:** This study found elevated GFAP levels in bvFTD compared to a large cohort of PPD, but NfL levels exhibited better performance in this distinction. These findings extend the literature on GFAP in bvFTD and build evidence for plasma NfL as a useful biomarker to assist with distinguishing bvFTD from PPD. Utilisation of NfL may improve timely and accurate diagnosis of bvFTD.

## INTRODUCTION

Timely and accurate diagnosis of behavioural variant frontotemporal dementia (bvFTD) remains a common challenge, especially when distinguishing from primary psychiatric conditions (Ducharme et al., 2020; Tsoukra et al., 2022). Blood biomarkers that could assist in routine clinical practice are much needed. There has been increasing research and interest in glial fibrillary acidic protein (GFAP), a marker of astrocyte dysfunction, as a diagnostic biomarker in neurodegenerative as well as psychiatric disorders (Abdelhak et al., 2022), and few studies have investigated the utility of both GFAP and other markers such as the marker of neuronal injury, neurofilament light chain (NfL).

Findings from studies investigating GFAP in bvFTD thus far have been mixed, with several studies finding elevated GFAP levels in bvFTD compared to controls (Benussi et al., 2020; Heller et al., 2020; Oeckl et al., 2022; Sanchez et al., 2024; Sarto et al., 2023; Zhu et al., 2021), while some have not (Baiardi et al., 2022; Chouliaras et al., 2022). One study compared GFAP levels in bvFTD to PPD, finding higher GFAP levels in bvFTD, diagnostic utility of GFAP to distinguish bvFTD from PPD, and additional diagnostic performance with combining GFAP and neurofilament light chain protein (NfL, a marker of neuronal injury) levels (Katisko et al., 2021). Regarding the GFAP in PPD literature, no studies have compared GFAP levels between a large mood and psychotic disorder cohorts. There is thus still a need for further understanding of GFAP and combination biomarkers in bvFTD and PPD, and their potential diagnostic and clinical utility.

In our previous study we investigated NfL finding strong diagnostic utility of this marker of neuronal injury to distinguish bvFTD from PPD (Eratne et al., 2024). The primary aim of this study was to follow up and investigate plasma GFAP levels in bvFTD compared to this large, diverse range of PPD (Aim 1), and the diagnostic utility of GFAP to distinguish bvFTD from PPD, compared to NfL and combination of GFAP and NfL (Aim 2). An exploratory aim was to explore differences in plasma GFAP levels between PPD and controls, and between individual PPD (Aim 3).

## METHODS

### Participant recruitment and data

This study was a follow up to our previous study that investigated NfL in four disease cohorts: bipolar affective disorder (BPAD), major depressive disorder (MDD), treatment-resistant schizophrenia (TRS), bvFTD, and a local control group (Eratne et al., 2024). Full details have been published previously but briefly: Cohort 1: patients with BPAD experiencing a bipolar depressive episode of at least moderate severity (n=121) (Berk et al., 2019). Cohort 2: patients with MDD experiencing a depressive episode of at least moderate severity (n=42) (Dean et al., 2014, 2017). Cohort 3: patients who were on clozapine with a diagnosis of treatment-resistant schizophrenia (TRS), defined as failure to respond to adequate trials of two or more antipsychotics (Bousman et al., 2019; Eratne et al., 2022; Mostaid et al., 2017). Cohort 4: patients with probable or definite bvFTD (n=22) based on comprehensive gold-standard expert multidisciplinary and multimodal investigations including structural and functional imaging (Ooi et al., 2022). For the control group, samples and data were pooled from healthy people (n=96) with no current or past psychiatric or neurological illness (healthy controls age-matched to TRS, and healthy parents and siblings of participants with TRS) (Eratne et al., 2022).

All studies that contributed data and samples to this study, had ethical approval at relevant Human Research Ethics Committees. All participants provided written informed consent prior to participation. This study, part of The Markers in Neuropsychiatric Disorders Study (The MiND Study, https://themindstudy.org), was approved by the Melbourne Health Human Research Ethics Committee (MH/HREC2020.142).

### Sample analysis

Plasma aliquots were stored at –80°C. Plasma GFAP and NfL levels were measured on Quanterix SR-X analysers using Simoa single assay kits, according to the manufacturer’s recommendations.

### Statistical analysis

Statistical analyses were performed using R version 4.3.2 (2023-10-31). Log_10_-transformed biomarker levels in different groups were compared using standardised bootstrapped generalised linear models (GLM), bootstrapped, with covariates age, sex and where available, weight. ROC curve analyses were performed to investigate diagnostic performance, with optimal biomarker cut-offs based on Youden’s J. Areas under the curve (AUC) for GFAP were compared to NfL from our previous study, and combination (sum) of GFAP and NfL, using bootstrapped DeLong test.

## RESULTS

The cohort included 245 patients with PPD (121 BPAD, 42 MDD, 82 TRS), 22 participants with bvFTD, 96 controls (Table 1). bvFTD was the oldest group (mean, M=66 years). MDD patients were older (M=55 years) than BPAD and TRS (mean 44 and 40 years, respectively).

**Table 1.** Details of study cohort. Data presented are mean (SD) or number (%) BPAD: bipolar affective disorder; bvFTD: behavioural variant frontotemporal dementia; MDD: major depressive disorder; TRS: treatment-resistant schizophrenia

|  | **BPAD**<br>**N=121** | **MDD**<br>**N=42** | **TRS**<br>**N=82** | **bvFTD**<br>**N=22** | **Control**<br>**N=96** |
| --- | --- | --- | --- | --- | --- |
| Age at sample (years) | 44.1 (12.2) | 55.0 (13.0) | 40.3 (9.32) | 65.7 (9.15) | 44.7 (14.4) |
| Sex female, n | 75 (62.0%) | 24 (57.1%) | 23 (28.0%) | 4 (18.2%) | 50 (52.1%) |
| Weight (kg) | 84.3 (20.9) | 84.6 (18.1) | 95.8 (24.9)<br>(n=71) | - | 77.3 (15.4)<br>(n=87) |
| Plasma GFAP (pg/mL) | 95.6 (49.3) | 105 (47.2) | 67.9 (53.0) | 273 (206) | 75.5 (46.9) |
| Log10GFAP | 1.93 (0.21) | 1.98 (0.19) | 1.73 (0.30) | 2.32 (0.33) | 1.80 (0.28) |

### Aim 1: Plasma GFAP levels in bvFTD and PPD

Plasma GFAP levels were significantly elevated in bvFTD (mean, M=273pg/mL), compared to all other groups (BPAD M=95.6pg/mL β=0.97 [0.49, 1.45], p<0.001; MDD M=105pg/mL β=1.05 [0.62, 1.53], p<0.001; TRS M=67.9pg/mL β=1.44 [0.99, 1.93], p<0.001; controls 75.5pg/mL, β=1.41 [0.92, 1.92], p<0.001), Table 1, Figure 1. Age and sex were significant covariates (p<0.001, Supplementary Material).

**Figure 1.**
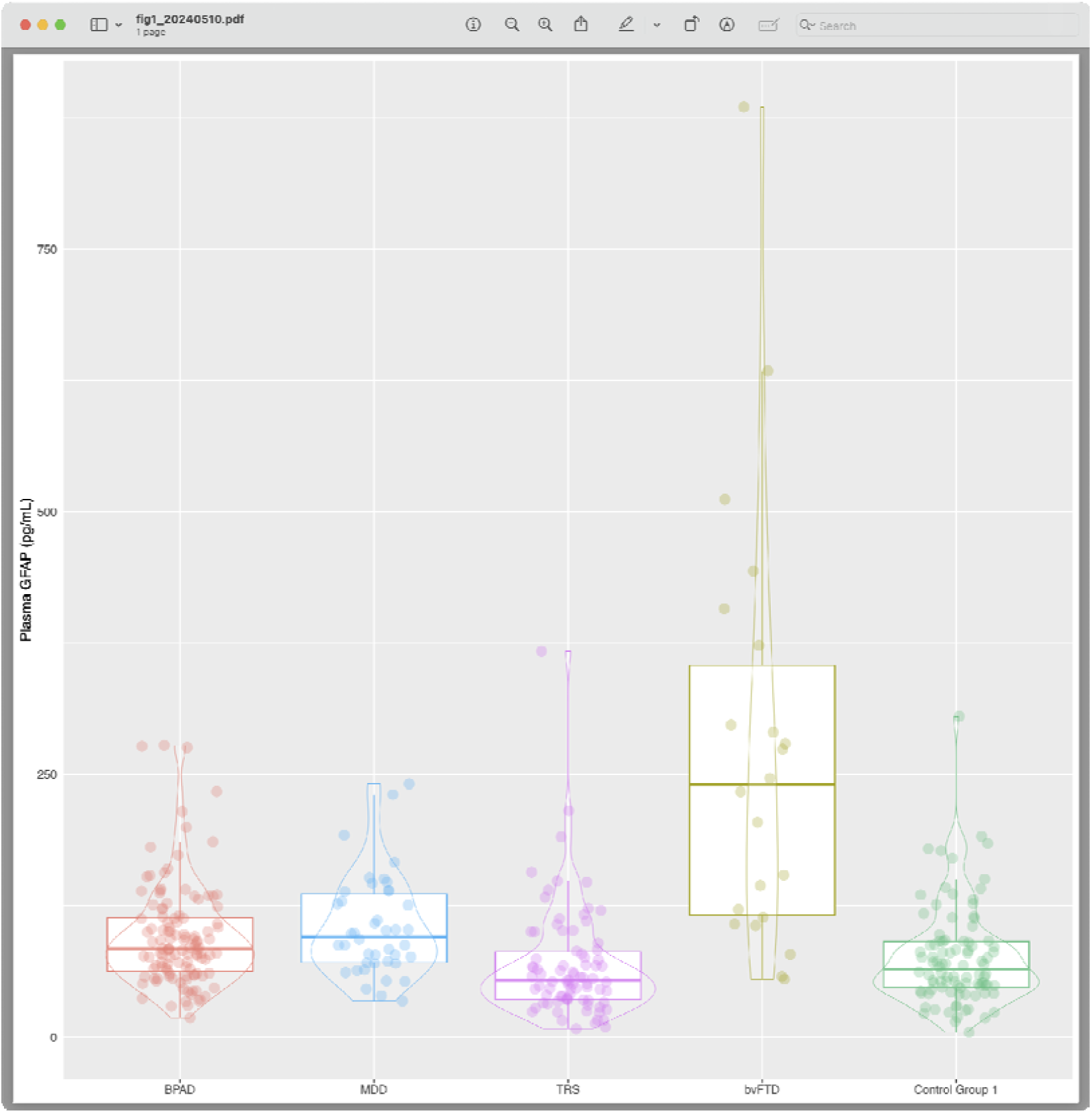
**Plasma GFAP levels in behavioural variant frontotemporal dementia, primary psychiatric disorders, and controls**. BPAD: bipolar affective disorder; bvFTD: behavioural variant frontotemporal dementia; MDD: major depressive disorder; TRS: treatment-resistant schizophrenia

### Aim 2: Diagnostic utility of plasma GFAP to distinguish bvFTD from PPD, compared to plasma NfL

GFAP distinguished bvFTD from all psychiatric disorders with an AUC of 0.86 [0.76, 0.95], Figure 2, with 105.1pg/mL optimal cut-off resulting in 73% specificity, 86% sensitivity.

**Figure 2.**
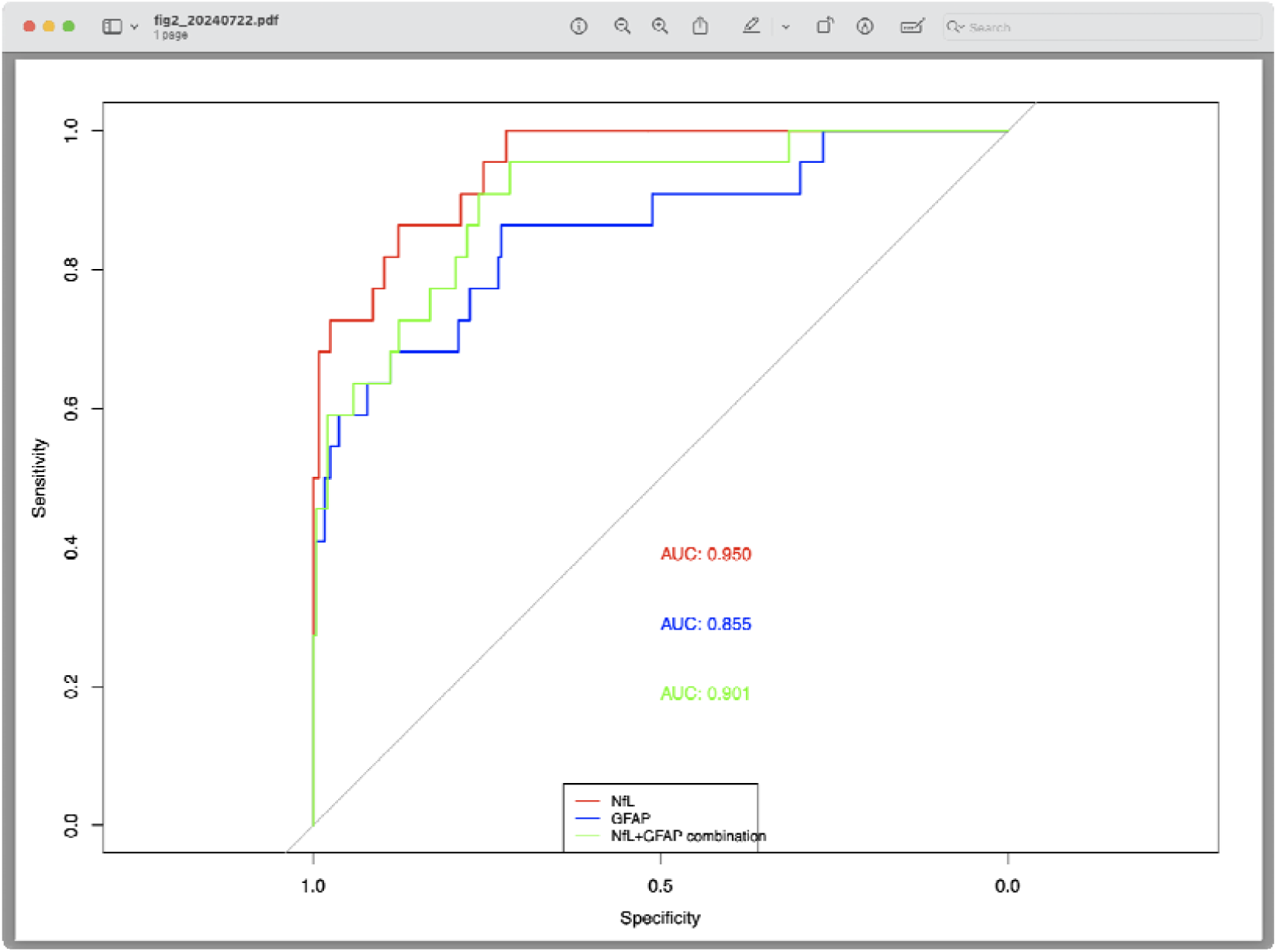
**ROC curve analysis of biomarkers to distinguish bvFTD from primary psychiatric disorders** AUC: area under the curve; BPAD: bipolar affective disorder; bvFTD: behavioural variant frontotemporal dementia; GFAP: glial fibrillary acidic protein; MDD: major depressive disorder; NfL: neurofilament light chain protein; TRS: treatment-resistant schizophrenia

We compared diagnostic performance of GFAP to that of NfL from our previous study on the same cohort (NfL AUC 0.95 [0.91, 0.99], cut-off 13.3pg/mL, 86% specificity, 88% sensitivity) (Eratne et al., 2024). NfL performed better than GFAP (statistically significant AUC difference 0.95 vs 0.86, p=0.02542). GFAP and NfL combination resulted in AUC 0.90 [0.83, 0.97], cut-off 114, 72% specificity, 95% sensitivity. While this GFAP+NfL combination AUC was better than GFAP alone (0.04726), it was still outperformed by NfL alone (p=0.0473).

### Aim 3: Plasma GFAP in primary psychiatric disorders compared to controls, and to each other

After adjusting for age, sex, and weight, plasma GFAP levels were higher in BPAD (β=0.58 [0.37, 0.78], p<0.001), and MDD (β=0.46 [0.19, 0.72], p<0.001), compared to controls (Supplementary Material). There was no evidence of difference between TRS and controls (β=0.09 [-0.25, 0.38], p=0.638). BPAD and MDD had higher levels than TRS (β=0.50 [0.22, 0.79], p=0.002 and β=0.38 [0.06, 0.73], p=0.014, respectively). Plasma GFAP did not demonstrate high diagnostic utility to differentiate: BPAD from controls (AUC 0.65 [0.57, 0.72], 54.2pg/mL cut-off, 41% specificity, 86% sensitivity), MDD from controls (AUC 0.71 [0.62, 0.80], 61.2pg/mL cut-off, 49% specificity, 88% sensitivity), BPAD from TRS (AUC 0.71 [0.64, 0.79], 68.9pg/mL cut-off, 69% specificity, 69% sensitivity), MDD from TRS (AUC 0.76 [0.68, 0.85], 69.2pg/mL cut-off, 69% specificity, 81% sensitivity).

## DISCUSSION

This follow-up study identified elevated GFAP levels in bvFTD compared to a large, diverse range of psychiatric disorders, and controls. GFAP nor a combination of GFAP and NfL, did not perform as well as NfL alone, to distinguish bvFTD from psychiatric disorders as described in our previous study (Eratne et al., 2024). This adds further evidence of biomarker abnormalities and astrocyte dysfunction in bvFTD, the superiority of plasma NfL to assist with distinguishing from psychiatric disorders, and extends the literature on GFAP in PPD, including the largest and most diverse PPD cohort to date.

While several studies have investigated GFAP in bvFTD compared to other neurodegenerative disorders and controls (Baiardi et al., 2022; Cousins et al., 2022; Sarto et al., 2023), to our knowledge, only one previous study has specifically investigated whether blood GFAP can distinguish frontotemporal lobar degeneration (FTLD, including bvFTD) from PPD (Katisko et al., 2021). That study had a larger number of bvFTD patients (n=66), but had smaller control (n=18) and PPD (n=44) groups, and less diverse PPD compared to the cohorts included in our study. Katisko et al. found elevated serum GFAP in bvFTD compared to PPD, diagnostic utility, and in addition found a combination of GFAP and NfL resulted in increased diagnostic performance to distinguish all FTLD from PPD. In contrast, we did not find any benefit of combining NfL and GFAP, with NfL as a single biomarker outperforming GFAP and the combination of NfL and GFAP, to distinguish bvFTD from PPD. This of relevance to potential clinical translation, given the practical ease of using a single biomarker over a combination.

Exploratory analyses found elevated GFAP levels in BPAD as well as MDD, compared to TRS and controls. This contrasts with our previous NfL findings, where we found elevated levels in BPAD compared to TRS and controls, but normal NfL levels in MDD (Eratne et al., 2024). While there have been previous studies on plasma GFAP in BPAD and MDD, to our knowledge, no other studies have compared plasma GFAP levels between BPAD, MDD, and TRS. One previous study had 45 participants with MDD, but only 11 BPAD and 9 schizophrenia (Steinacker et al., 2021). We found higher GFAP levels in MDD and similar AUC for MDD vs controls (Supplementary Material), but in contrast, we also found elevated levels in BPAD. These differential biomarker changes in BPAD compared to MDD could potentially highlight differing physiological abnormalities in different disorders, for example possibly in BPAD there being both a degree of neuroinflammation and reactive astrogliosis (elevated GFAP) and neuronal injury (elevated NfL), whereas in MDD neuroinflammation and reactive astrogliosis (elevated GFAP), but without neuronal injury (NfL not elevated). Studies to investigate these findings in detail, especially associations with important clinical covariates, are currently underway.

Limitations include the relatively small number of bvFTD, lack of longitudinal follow up and definitive (e.g. genetic) confirmation, and limitations inherent to a retrospective design. Replication is needed in large, clinical cohorts, including neuroimaging and broad clinical covariates, and serial levels.

In summary, we found elevated plasma GFAP levels in bvFTD compared to a large and diverse cohort of PPD, diagnostic utility of GFAP levels to distinguish bvFTD from PPD, while reinforcing the superiority of NfL in this distinction. These findings build important evidence for biomarker changes and utility of different biomarkers in this often-challenging clinical distinction.

## Data Availability

All data produced in the present study are available upon reasonable request to the authors

## ACKNOWLEDGEMENTS AND FUNDING SOURCES

Finally, the authors would like to thank all the patients and their families for their participation.

The corresponding author had full access to all the data in the studscy and had final responsibility for the decision to submit for publication.

The Treatment Resistant Schizophrenia (TRS) biobank was established by IE, CP, CB as part of a major Australian Department of Industry Co-operative Research Centre (CRC) grant – https://researchdata.ands.org.au/treatment-resistant-schizophrenia-biobank/1325206

C Pantelis was supported by a National Health and Medical Research Council (NHMRC) L3 Investigator Grant (1196508) and NHMRC Program Grant (ID: 1150083).

HZ is a Wallenberg Scholar and a Distinguished Professor at the Swedish Research Council supported by grants from the Swedish Research Council (#2023-00356; #2022-01018 and #2019-02397), the European Union’s Horizon Europe research and innovation programme under grant agreement No 101053962, and Swedish State Support for Clinical Research (#ALFGBG-71320).

## DECLARATION OF INTERESTS AND FINANCIAL DISCLOSURES

All the authors have nothing to disclose. AB serves on the editorial boards of Neurology, Stroke and International Journal of Stroke and Scientific Advisory Boards for Eisai, Eli Lilly, Biogen and Novo Nordisk. HZ has served at scientific advisory boards and/or as a consultant for Abbvie, Acumen, Alector, Alzinova, ALZPath, Amylyx, Annexon, Apellis, Artery Therapeutics, AZTherapies, Cognito Therapeutics, CogRx, Denali, Eisai, LabCorp, Merry Life, Nervgen, Novo Nordisk, Optoceutics, Passage Bio, Pinteon Therapeutics, Prothena, Red Abbey Labs, reMYND, Roche, Samumed, Siemens Healthineers, Triplet Therapeutics, and Wave, has given lectures in symposia sponsored by Alzecure, Biogen, Cellectricon, Fujirebio, Lilly, Novo Nordisk, and Roche, and is a co-founder of Brain Biomarker Solutions in Gothenburg AB (BBS), which is a part of the GU Ventures Incubator Program (outside submitted work).

### Statistical analysis conducted by

Dr Dhamidhu Eratne

## Appendix: collaborators/contributors

On behalf of others in The MiND Study Group:

**On behalf of others in The MiND Study Group:**
| **Name** | **Affiliations (separated by semi colon)** |
| --- | --- |
| Hannah Dobson | Psychiatry, Alfred Health; Neuropsychiatry, Royal Melbourne Hospital |
| Rosie Watson | Population Health and Immunity Division, The Walter and Eliza Hall Institute of Medical Research, Parkville, Australia;<br>Department of Aged Care and Medicine, The Royal Melbourne Hospital, University of Melbourne, Parkville, Australia |
| Nawaf Yassi | Population Health and Immunity Division, The Walter and Eliza Hall Institute of Medical Research, Parkville, Australia;<br>Department of Medicine and Neurology, Melbourne Brain Centre at the Royal Melbourne Hospital, University of Melbourne, Parkville, Australia |
| Terence J. O'Brien | Department of Neuroscience, The Central Clinical School, Monash University |
| Patrick Kwan | Department of Neuroscience, The Central Clinical School, Monash University |
| Christiane Stehmann | Australian National CJD Registry, The Florey Institute, Melbourne |
| Christopher Fowler | The Florey Institute of Neuroscience and Mental Health, The University of Melbourne, Parkville, VIC |
| Sarah Farrand | Neuropsychiatry, Royal Melbourne Hospital; Melbourne Neuropsychiatry Centre & Department of Psychiatry, University of Melbourne |
| Michael Keem | Neuropsychiatry, Royal Melbourne Hospital |
| Cath Kaylor-Hughes | Integrated Mental Health Team<br>Dept of General Practice<br>University of Melbourne, 780 Elizabeth St, Melbourne, 3010 |
| Richard Kanaan | Dept of Psychiatry, University of Melbourne, Austin Health, Heidelberg, VIC 3084 |
| Piero Perucca | Department of Medicine, Austin Health, The University of Melbourne; Comprehensive Epilepsy Program, Austin Health; Department of Neuroscience, Central Clinical School, Monash University; Department of Neurology, The Royal Melbourne Hospital; Department of Neurology, Alfred Health, Melbourne, VIC, Australia |
| Rashida Ali | Alfred Health; Monash University |
| Anna King | The Wicking Dementia Centre, Tasmania |
| Jane Gunn | Department of General Practice, The University of Melbourne |
| Tianxin Pan | Health Economics Unit \| Centre for Health Policy \| Melbourne School of Population and Global Health, The University of Melbourne |
| Ilias Goranitis | Health Economics Unit \| Centre for Health Policy \| Melbourne School of Population and Global Health, The University of Melbourne |
| Steve Simpson-Yap | CORe, The Royal Melbourne Hospital, The University of Melbourne, Melbourne, VIC, Australia |
| Tomas Kalincik | CORe, The Royal Melbourne Hospital, The University of Melbourne, Melbourne, VIC, Australia; Neuroimmunology Centre, Department of Neurology, The Royal Melbourne Hospital, Melbourne, VIC, Australia |

## SUPPLEMENTARY MATERIAL

### Results of generalised linear model comparing bvFTD (reference) to other disorders, adjusting for age and sex

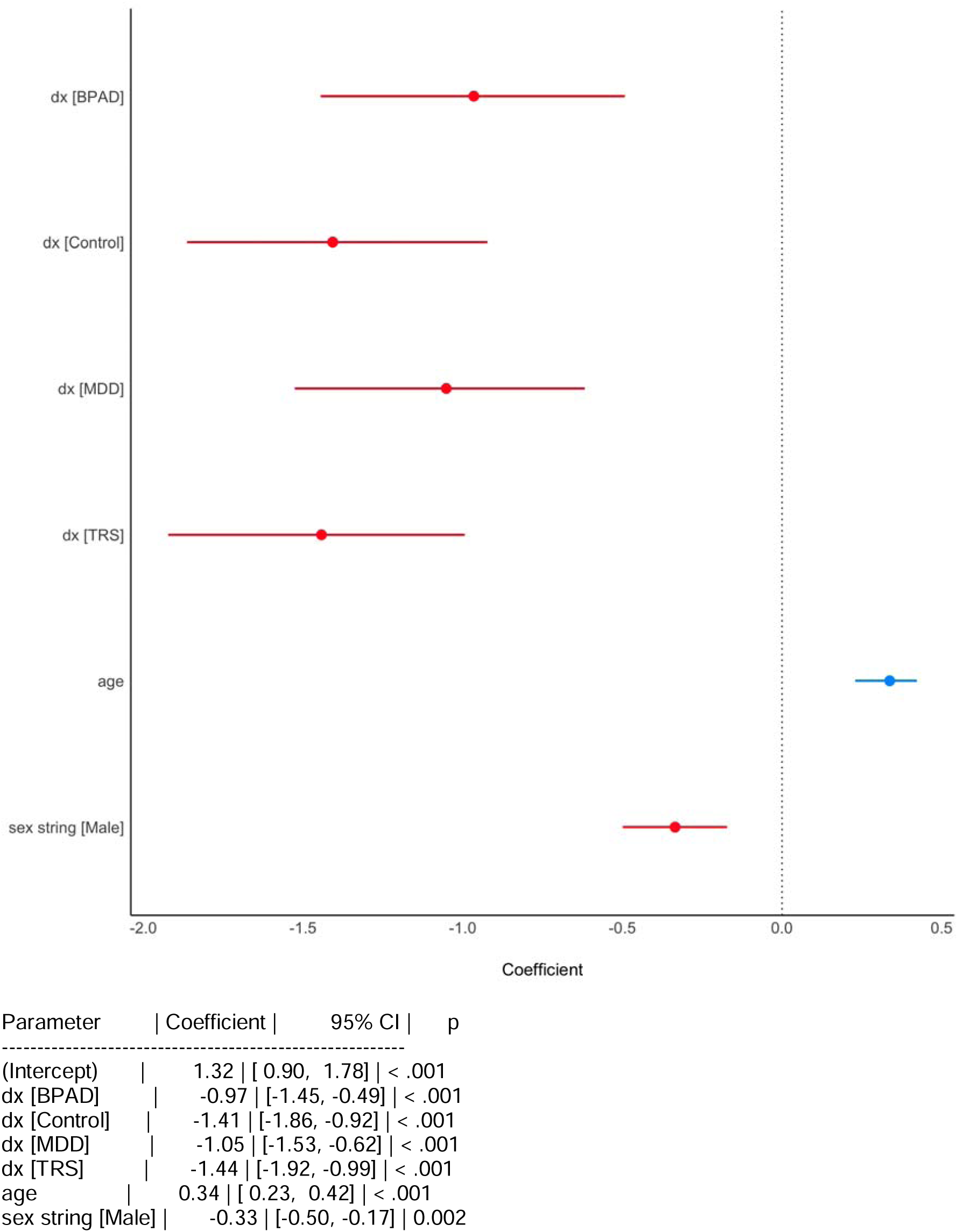

### Results of generalised linear model comparing PPD to Controls (reference), adjusting for age, sex, and weight

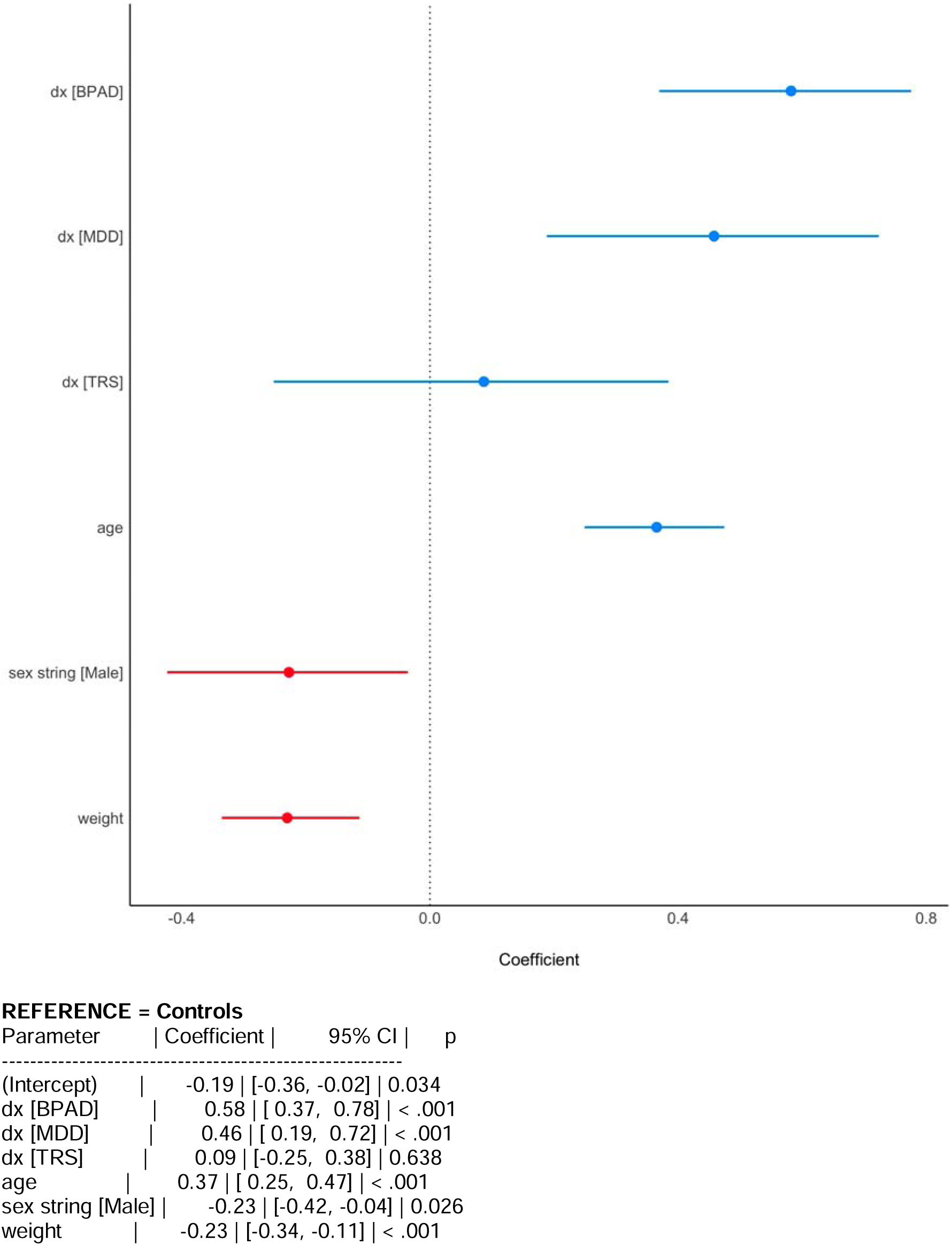

### Results of generalised linear model comparing PPDs and controls to TRS (reference), adjusting for age, sex, and weight

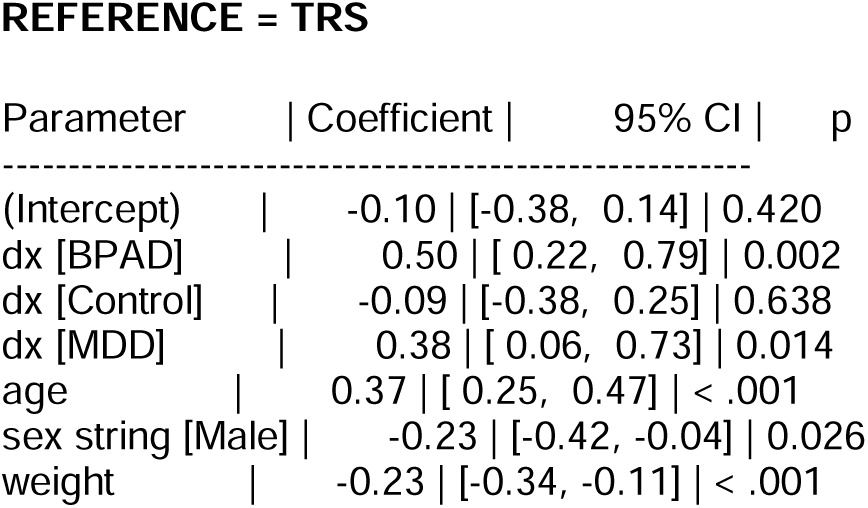

### GFAP to distinguish PPD from controls and from each other

Plasma GFAP did not demonstrate high diagnostic utility to differentiate: BPAD from controls (AUC 0.65 [0.57, 0.72], 54.2pg/mL cut-off, 41% specificity, 86% sensitivity), MDD from controls (AUC 0.71 [0.62, 0.80], 61.2pg/mL cut-off, 49% specificity, 88% sensitivity), BPAD from TRS (AUC 0.71 [0.64, 0.79], 68.9pg/mL cut-off, 69% specificity, 69% sensitivity), MDD from TRS (AUC 0.76 [0.68, 0.85], 69.2pg/mL cut-off, 69% specificity, 81% sensitivity).

## Notes

### Competing Interest Statement

The authors have declared no competing interest.

### Author Declarations

All participants provided written informed consent prior to participation. This study was approved by the Melbourne Health Human Research Ethics Committee (MH/HREC2020.142).

